# Assessment of Glucose Metabolism *In Vivo* in the Human Frontal Lobe Using Interleaved ^1^H and ^13^C MRS at 7T: Toward Clinical Translation

**DOI:** 10.64898/2026.07.25.26358922

**Authors:** Ying Xiao, Daniel Wenz, Indrit Bègue, Patric Hagmann, João M N Duarte, Loan Mattera, Nathalie Philippe, Antonia Kaiser, Katarzyna Pierzchala, André Döring, Mark Widmaier, Kim Q. Do, Rolf Gruetter, Dimitrios C. Karampinos, Lijing Xin

**Author notes:** **Corresponding author:** Lijing Xin, Institute of Physics, School of Basic Sciences, CIBM MRI EPFL CH F1 532 Station 6, 1015 Lausanne, Switzerland.

## Abstract

**Background:** Mitochondrial dysfunction and abnormal cerebral energy metabolism are implicated in many neuropsychiatric and neurodegenerative disorders. ^13^C magnetic resonance spectroscopy (MRS), combined with ^13^C-labeled substrate infusion, offers a non-ionizing, minimally invasive method for assessing fluxes through the main cerebral energy metabolism pathways. However, its human application at 7 T has not been fully established, especially within the frontal lobe.

**Purpose:** To explore a clinically translatable interleaved ^1^H/^13^C MRS protocol for quantification of cerebral glucose uptake and downstream metabolism at 7 T, and to estimate the tricarboxylic acid (TCA) cycle flux (V_TCA_) for validation.

**Study Type:** Prospective.

**Population:** Three young healthy volunteers.

**Field Strength/Sequence:** 7T; ACE-STEAM (indirect ^1^H-[^13^C]) and ISIS-DEPT (direct ^13^C-[^1^H]).

**Assessment:** ACE-STEAM and ISIS-DEPT were applied to acquire the time-resolved spectra in the frontal lobe. ^13^C-labeled glucose, glutamate, and glutamine fractional enrichment time courses were quantified to estimate V_TCA_ through the one-compartment model.

**Statistical Tests:** The relative estimated fitting uncertainties (EFUs) were reported for the processed spectra. Nonlinear least squares minimization was used for flux fitting of ^13^C traces. Uncertainty of the estimated metabolic fluxes was evaluated using Monte-Carlo simulations.

**Results:** [1-^13^C]-glucose (GlcC1) was detected immediately on ^13^C MR spectra, followed by ^13^C-labeled GluH4 and GlnH4 and then GlxH3 can be quantified on ^1^H MR spectra. End-of-infusion mean enrichments were 17% (GluH4), 13% (GlnH4), and 7% (GlxH3). Brain glucose concentration ranged 1.86-2.94 mM, with 61% of the mean enrichment in C1. Group-average V_TCA_ was 0.66 ± 0.07 μmol/g/min.

**Data Conclusion:** This interleaved ^1^H/^13^C MRS protocol enables minimally invasive quantification of cerebral metabolic fluxes, may provide a useful framework for investigating neuropsychiatric and neurodegenerative diseases at 7 T.

**Evidence Level:** 1.

**Technical Efficacy:** Stage 1.

## Introduction

Energy metabolism is crucial for maintaining the structural and functional integrity of the brain.^1^ As the most energy-demanding organ, the brain relies on efficient metabolic processes to sustain complex activities, including synaptic transmission, plasticity, and redox homeostasis, which are critical for cognitive functions. Impaired energy metabolism and mitochondrial dysfunction in the frontal cortex are increasingly linked to cognitive decline in neurodegenerative, neuropsychiatric disorders, and aging.^2,3,4^ ^13^C magnetic resonance spectroscopy (MRS), combined with the administration of ^13^C-labeled substrates such as [1-^13^C] glucose, provides a powerful approach to quantitatively investigate cerebral metabolic processes by tracing the incorporation of ^13^C into downstream metabolites, namely glutamate and glutamine.^5^ This technique has demonstrated outstanding utility and has been widely applied in preclinical studies.^6,7,8,9,10,11^

With the growing availability of 7 T MRI systems, implementing ^13^C MRS at this field strength offers clear advantages in spectral resolution and sensitivity. Earlier human studies^12,13,14,15,16,17,18^ performed at lower fields (1.5-4 T) have utilized direct ^13^C detection to quantify tricarboxylic acid (TCA) cycle fluxes and neurotransmitter cycling; but these approaches typically rely on broadband proton decoupling to enhance sensitivity and simplify spectra. However, such decoupling becomes difficult to implement at 7 T because of strict SAR limitations^19^, a challenge that is especially pronounced in the frontal lobe, where proximity to the eyes reduces the tissue’s ability to dissipate RF-induced heating.^20^ Proton-Observed, Carbon-Edited (POCE)^21,22,23,24,25^ methods have shown that ^13^C labeling curves can be assessed indirectly through ^1^H detection; however, indirect detection cannot fully benefit from the broader spectral dispersion of ^13^C, resulting in more crowded spectra and reduced ability to resolve individual metabolites compared with direct ^13^C MRS.

Translating these methods into clinical practice also presents several other challenges.^26^ Many of the previous studies inferred brain glucose dynamics from plasma to quantify metabolic fluxes. These studies typically modeled the input function using Michaelis–Menten kinetics^27^ under the assumption of saturated glucose transport. As a result, most previous studies required intravenous infusion of high-dose (∼0.7 g/kg), hyperosmolar (20%) glucose solution^19,28,29^, which often exceeded standard peripheral infusion tolerances. Such solutions increase the risk of venous irritation, phlebitis, and fluid infiltration.^30^ These safety concerns, the requirements of blood sampling, alongside high material costs, thereby limit the feasibility and broader clinical applicability of ^13^C MRS.

The technical and safety challenges mentioned above have limited the broader clinical implementation of ^13^C MRS for the human frontal lobe at 7 T. In this study, we introduce an integrated framework designed to overcome these constraints and establish a clinically feasible protocol. We developed a multi-channel RF surface coil optimized for frontal-lobe coverage. To ensure safety and participant comfort, we employed a moderate glucose infusion protocol. We implemented acquisition and quantification strategies that substantially reduce SAR without the need for proton or carbon decoupling. The MRS acquisition integrates interleaved indirect and direct ^13^C MRS acquisitions (ACE-STEAM^6^ and ISIS-DEPT^8^), enabling direct measurement of brain glucose fractional enrichment as the metabolic input function. The proposed design eliminates the need for blood sampling while still providing sufficient data for estimating metabolic fluxes, namely V_TCA_. Overall, the goal of this work was to establish a robust, less-invasive, and patient-friendly method for frontal-lobe ^13^C MRS at 7 T, and to provide a practical framework for investigating cerebral glucose metabolism that could be later translated to clinical settings.

## Methods

### Participants and glucose infusion protocol

Three healthy volunteers (two females and one male, aged 31-32 years, body mass index of 19.5-22.9) were recruited for the study. All procedures were approved by the local ethics committee, and written informed consent was obtained from all participants. Fasting blood glucose was confirmed normal by finger-prick screening before participation. Participants were monitored for two hours post-scan to ensure safety.

Participants were instructed to fast overnight (>9 hours) before the scan. An intravenous catheter was inserted into a vein in the lower arm for the infusion of the ^13^C-labeled glucose solution. A second catheter was placed in the opposite arm for periodic blood sampling to measure plasma glucose for comparison with MRS-measured data.

An infusion solution of 10% (w/v) made by 99% enriched [1-^13^C]-glucose (Sigma-Aldrich Chemie GmbH, OH, USA). The 10% (w/v) [1-^13^C]-glucose solution was administered intravenously using an automated syringe pump over approximately 60 minutes with a total dose of 0.5 g/kg body weight using a two-phase protocol: the initial bolus phase (first 14 minutes), consisting of half the total dose delivered at three rates aiming to raise glucose by 7 mM, produced a rapid rise in the glucose level. This was followed by a slower maintenance infusion with a constant rate to sustain stable cerebral ^13^C enrichment of glucose (protocol detailed in Supplementary Material Section 1).

### RF coil

Participants were positioned supine in a 7T MRI scanner (Terra.X 7T, Siemens, Erlangen, Germany) using a custom-built dual-tuned RF surface coil (3-channel ^1^H / 2-channel ^13^C; Figure 1 (a)). The coil was aligned with the forehead. Electromagnetic simulations using an FDTD solver in Sim4Life (ZMT Zurich MedTech, Switzerland) were performed to evaluate local SAR (10 g averaged) and whole-body average SAR for the ^1^H and ^13^C RF coils under realistic loading conditions, and all RF exposures were confirmed to remain within IEC 60601-2-33 Normal Operating Mode safety limits for human 7 T operation. Coil design and bench tests (E5071C, Agilent Technologies) are provided in Supplementary Material Section 2.

**Figure 1:**
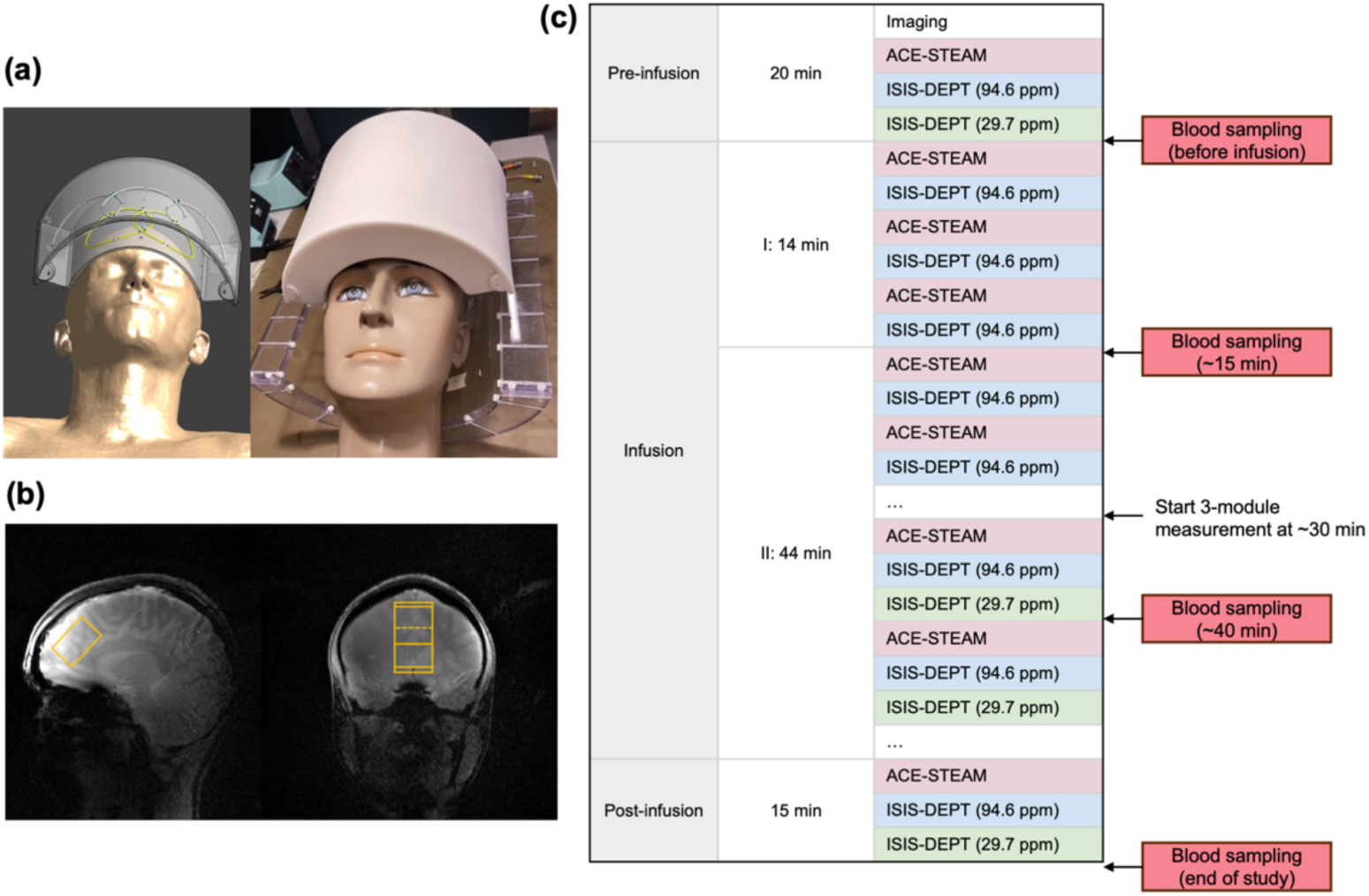
Experimental setup and example design: (a) Illustration of the dual-tuned 3-channel ^1^H / 2-channel ^13^C surface coil. Left: simulated coil placement on the Duke human model using Sim4Life.Right: Photograph of the coil setup. All loops are enclosed in the casing designed for frontal lobe coverage, mounted on two adjustable arms and a transparent base to facilitate precise positioning. (b) voxel (2.5 × 2.8 × 4.0 cm^3^) localization in the frontal lobe. (c) Example interleaved MRS acquisition scheme. The session began with around 20 minutes allocated to preparation scans, including 3D anatomical imaging for voxel localization, shimming, and baseline MRS measurements (before infusion). Intravenous infusion of 10% [1-^13^C]-glucose solution (total dose: 0.5 g/kg body weight) included a priming bolus over the first 14 minutes (50% of the dose, delivered at three rates), followed by a slower maintenance infusion. The ^1^H and ^13^C MRS were interleaved to form one complete acquisition block: (1) ACE-STEAM, (2) ISIS-DEPT carried at 94.6 ppm, and (3) ISIS-DEPT carried at 29.7 ppm conducted after 30 minutes. After completion of the infusion (at ∼60 minutes), one final block of MRS acquisition was performed. Venous blood was sampled at baseline (pre-infusion), then about every 20 minutes during the infusion phase, and once more at the end of the full experiment.

### MR protocol

The frontal lobe MRS voxels for ^1^H (2.8 × 2.5 × 4.0 cm^3^; Figure 1(b)) and ^13^C (3.0 × 3.0 × 4.0 cm^3^) were aligned at the same center position, with placement based on a 3D gradient echo image (TR/TE = 8.60/3.69 ms, flip angle = 90°, matrix = 512 × 512, FOV = 25 × 25 cm^2^, slice thickness = 5 mm). The B₀ magnetic field homogeneity within the voxel was optimized using the vendor shimming method (“GRE-brain”) and FAST(EST)MAP^31^. ^13^C frequency calibration was adjusted using a 1-cm diameter sphere containing 99% [1-^13^C] formic acid, positioned at the coil center.

MRS data were acquired dynamically by an interleaved protocol that alternated between direct and indirect single-voxel spectroscopy sequences:

1. ACE-STEAM (Adiabatic Carbon-Edited Stimulated Echo Acquisition Mode)^6^ combined with interleaved Outer Volume Saturation (OVS) and variable pulse power and optimized relaxation delay (VAPOR) water suppression^32^ was used for scanning indirect ^1^H-[^13^C] MR spectra, with parameters TE/TM/TR = 7.9/35/4000 ms, 4096 data points, and a spectral bandwidth of 8 kHz.
2. ISIS-DEPT (Image-Selected In Vivo Spectroscopy combined with Distortionless Enhancement by Polarization Transfer)^8^ was employed for acquiring the direct ^13^C-[^1^H] MR spectra. The receiver was centered at 59.9 ppm. To target metabolites of interest, the ^13^C RF pulse carrier was set to 94.6 ppm for [1-^13^C] glucose (GlcC1) and to 29.7 ppm for [4-^13^C] or [3-^13^C] glutamate and glutamine (GlxC4 or GlxC3). The flip angle of the last pulse applied on the ^1^H frequency (θ) was adjusted accordingly: 90° for C1 of glucose detection and 45° for C3 and C4 of glutamate and glutamine. Other acquisition parameters: TR = 4s, 4096 data points, and spectral bandwidth = 20 kHz.

Two measurement modules were interleaved to form each acquisition block during the first 30 minutes: (1) ACE-STEAM, and (2) ISIS-DEPT at 94.6 ppm. After 30 minutes, a third module, ISIS-DEPT at 29.7 ppm, was added, followed by a final 10-minute ISIS-DEPT acquisition at 29.7 ppm. Each module (except the last one) comprised 32 transients (∼2 minutes per module). No ^1^H or ^13^C decoupling was applied. Pulse sequences were detailed in Supplementary Material Section 3.

The scan included ∼20 minutes for anatomical imaging, voxel placing, shimming, and baseline scans, and ∼75 minutes of the MRS acquisition during and after [1-^13^C]-glucose infusion (Figure 1(c)). Unsuppressed water reference spectra were acquired at baseline and after each blood sampling using ACE-STEAM with identical parameters without water suppression and OVS, for coil combination and linewidth measurement.

### Venous blood sampling and processing

For venous blood sampling, the scanner bed was briefly moved out without interrupting the infusion, after which participants were repositioned for continued scanning. Blood was drawn from the contralateral antecubital vein, with approximately 4 mL collected for each sample. Blood glucose level was measured immediately using a glucose meter (Axapharm, Baar, Switzerland). The remaining sample was centrifuged at 4 °C to separate plasma, which was stored at −80 °C until analysis. Plasma ^13^C isotopic enrichment of glucose was quantified using liquid chromatography-high-resolution mass spectrometry (LC-HRMS).

### Spectral processing

#### 1H and ^1^H-[^13^C] MR spectra

All spectral processing was performed using in-house MATLAB scripts (MathWorks, Natick, MA). The time-domain data were then truncated to 1024 points and zero-filled to 4096 after coil combination, frequency alignment, and water residual removal. The resulting spectra were analyzed using two approaches:

1. Difference spectra: subtracting the transients with the editing pulse on and off provided the ^1^H signals from ^13^C-coupled metabolites (e.g., GluH4, GlnH4, and GlxH3);
2. Sum spectra: summing the transients with the editing pulse on with those off provided unlabeled signals of metabolites.

### 13C-[1H] MR spectra

The raw data were first truncated to 1024 datapoints, zero-filled to 4096, and then denoised using the Marchenko–Pastur PCA (MP-PCA) method^33^. An exponential line-broadening filter of 5 Hz was applied. The GlcC1 spectrum acquired at the end of the infusion was used as the reference for ^13^C coil combination, phase correction, and frequency alignment of all ISIS-DEPT spectra.

The MRSinMRS checklist^34^ is included in Supplementary Material Section 4.

### Metabolite quantification

Metabolite quantification was performed using LCModel^35^ (Version 6.3-1N) with basis sets simulated using the density-matrix formalism. Spin parameters, including the chemical shifts and J_HH_-coupling constants, were taken from the literature^36,37^. J_CH_ coupling constants for the relevant resonances were experimentally determined from the analysis of multiplet structures observed in the ^13^C MR spectra, with values summarized in the Supplementary Material Section 5.

The basis sets used to fit the spectra include the following metabolites:

1. ACE-STEAM sum spectra (^1^H resonances): aspartate (Asp), creatine (Cr), phosphocreatine (PCr), glucose (Glc), glutamine (Gln), glutamate (Glu), glutathione (GSH), glycerophosphorylcholine (GPC), phosphorylcholine (PCho), myo-inositol (Ins), lactate (Lac), N-acetylaspartate (NAA), N-acetylaspartylglutamate (NAAG), phosphorylethanolamine (PE), taurine (Tau), γ-aminobutyric acid (GABA), and experimentally measured macromolecules (MM);
2. ACE-STEAM difference spectra (^13^C-coupled ^1^H resonances): [4-^13^C]-glutamate-H4 (GluH4), [4-^13^C]-glutamine-H4 (GlnH4), [3-^13^C]-glutamate-H3 (GluH3), and [3-^13^C]-glutamine-H3 (GlnH3);
3. ISIS-DEPT spectra carried at 94.6 ppm: [1-^13^C]-α-D-glucose (α-GlcC1) and [1-^13^C]-β-D-glucose (β-GlcC1);
4. ISIS-DEPT spectra carried at 29.7 ppm: [4-^13^C]-glutamate (GluC4), [4-^13^C]-glutamine (GlnC4), [3-^13^C]-glutamate (GluC3), and [3-^13^C]-glutamine (GlnC3).

All metabolite concentrations reported in millimolar (mM) in this study were scaled relative to total creatine (tCr) determined from baseline ACE-STEAM sum spectra. A fixed concentration of 8 mM tCr was assumed for all quantifications^38^.

The isotopic fractional enrichments (FEs) of C3 and C4 of glutamate and glutamine were calculated as the ratio of the ^13^C-labeled resonances (GluH3, GluH4, GlnH3, and GlnH4 measured from ACE-STEAM difference spectra) to the corresponding total glutamate and glutamine pool (obtained from ACE-STEAM sum spectra acquired at the baseline). The FE of GlcC1 was calculated using the GlcC1 concentration measured from ISIS-DEPT and the total glucose concentration quantified from ACE-STEAM sum spectra within the same acquisition block.

The concentration of GlcC1 was calculated from the β-anomeric using GlcC1 = β-GlcC1/0.64, considering that β-D-glucose represents 64% of the total D-glucose pool. The absolute concentration of β-GlcC1 was determined by scaling against the GluH4 concentration measured at the final time point using ACE-STEAM difference spectra and GluC4 from ISIS-DEPT (29.7 ppm), according to: 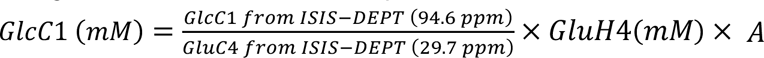, where A is the relative signal enhancement factor between these two resonances, determined from a self-built phantom containing 100 mM glutamate and 100 mM glucose, using the same acquisition sequence.

### Metabolic fluxes estimation

Cerebral metabolic fluxes were quantified using a one-compartment kinetic model based on the dynamic ^13^C label time courses.^15,39,40^ The model was implemented as a system of coupled ODEs describing the dynamic ^13^C label of key intermediates (Supplementary Material Section 6).

Subject-specific and group-average input functions were generated by fitting the GlcC1 FE data to an exponential model^40^: *FE_GlcC1_(t) = a × (1 − e^bt^*). Parameter *a* represents the expected steady-state FE level, while parameter *b* characterizes the rate approached to this plateau. Using the group-average input, the measured FE time courses of GluC4, GlnC4, and GlxC3 were simultaneously fitted to determine the fluxes V_TCA_, V_X_, and V_GLN_, via the nonlinear least-squares optimization. Uncertainty in fluxes was assessed using Monte-Carlo simulations (100 iterations) that incorporated variability in the estimated FEs of GluC4, GlnC4, and GlxC3 across participants, reported as the standard deviation. Individual V_TCA_ assessment for each participant was performed by fixing V_X_ and V_GLN_ to the group-mean values. Uncertainty was assessed with Monte Carlo simulations (100 iterations per participant), incorporating variability in the quantified FEs.

## Results

### Time-resolved MR spectra

Time-resolved ^1^H and ^13^C MR spectra are shown in Figure 2. High-quality spectra were obtained, showing well-resolved ^13^C-coupled ^1^H resonances (GluH4, GlnH4, GluH3/GlnH3) and ^13^C resonances of glucose (GlcC1), glutamate (GluC4, GluC3), and glutamine (GlnC4, GlnC3). The ACE-STEAM sum spectra were highly stable with the water linewidth of 13.1 ± 0.6 Hz across all studies. The difference spectra revealed dynamic incorporation of [1-^13^C]-glucose into glutamate and glutamine over infusion. GluH4 doublets (2.11 and 2.56 ppm) appeared ∼20 minutes after infusion onset, followed by GlnH4 (2.24 and 2.66 ppm) and GlxH3 (∼30 minutes), reflecting sequential labeling in the TCA and glutamate-glutamine cycles, as shown in Figure 2(a) and (b).

**Figure 2:**
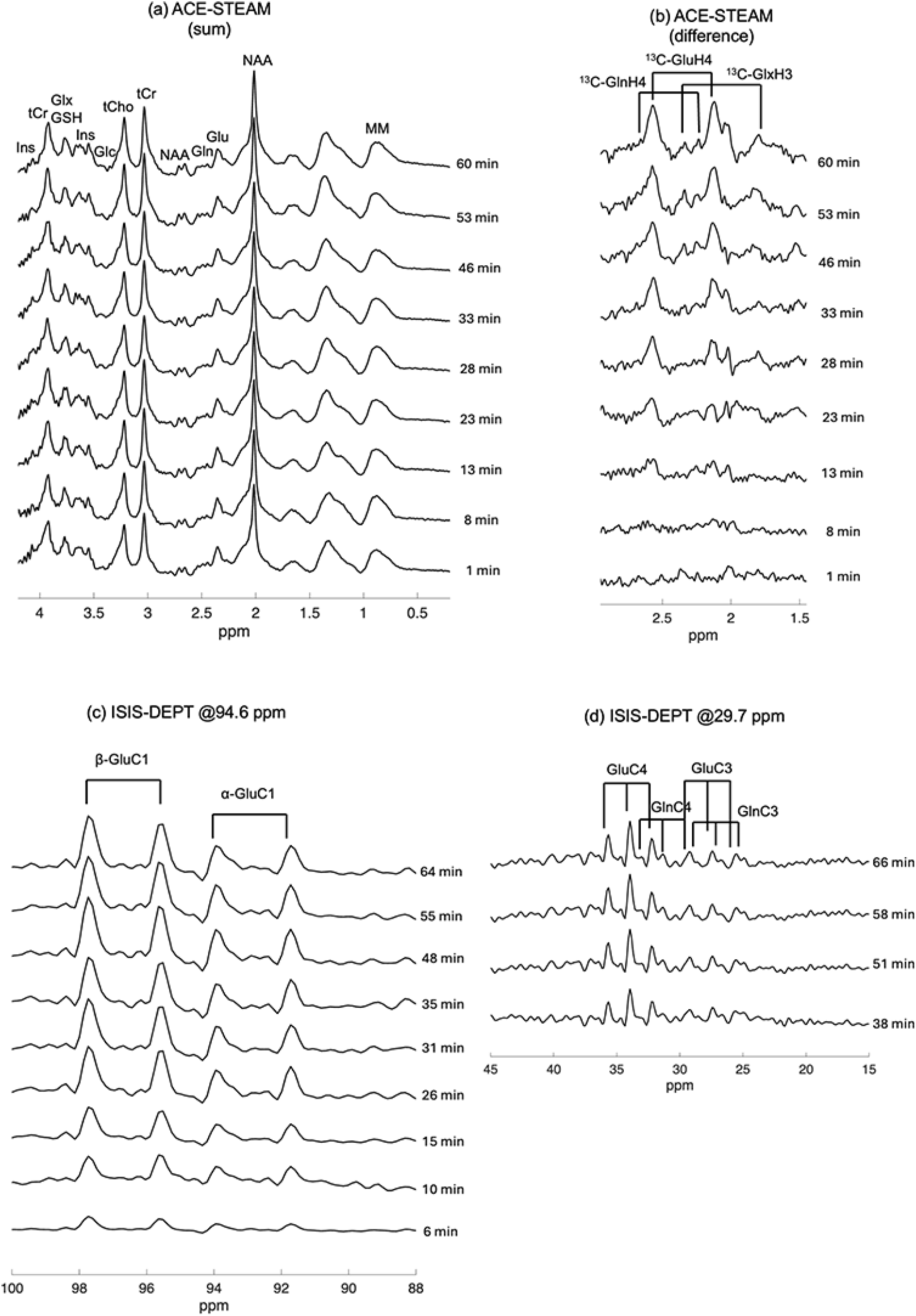
The time-resolved MR spectra. (a,b): ACE-STEAM (TE/TR = 7.9/4000 ms, 32 averages, spectral bandwidth = 8 kHz, data points = 4096, 4 Hz of Lorentzian linebroadening for difference spectra); (c,d) ISIS-DEPT (TR = 7.9/4000 ms, 32 averages, spectral bandwidth = 20 kHz, data points = 4096, 5 Hz of Lorentzian linebroadening) with alternating transmit offset. Each sequence was acquired in 2-minute blocks (32 averages).

Figure 3 illustrates representative spectra acquired using ACE-STEAM and ISIS-DEPT, along with corresponding spectral decompositions. Figure 3(a) shows the ACE-STEAM sum spectrum, processed with macromolecule and baseline removal, achieved by subtracting the fitted components obtained from the LCModel fitting results. Within the 2.9-4.0 ppm region, the glucose can be separated, and resonances were also resolved into key metabolite contributions, including tCr, Gln, Glu, GSH, tCho, mI, PE, and Tau. Figure 3(b) presents the ACE-STEAM difference spectrum, which detects ^13^C-coupled ^1^H signals. Distinct doublets corresponding to GluH4, GlnH4, and GlxH3 were resolved. Figure 3(c) displays a clear separation of the two glucose doublets (α-GlcC1 and β-GlcC1) from 91.8 to 97.6 ppm, reflecting cerebral uptake of [1-^13^C]-glucose. Figure 3(d) shows that the ^13^C spectrum from 25 to 36 ppm can resolve four distinct triplets corresponding to GluC4, GlnC4, GluC3, and GlnC3. This separation enables differentiation of glutamate and glutamine at C3 carbon positions.

**Figure 3:**
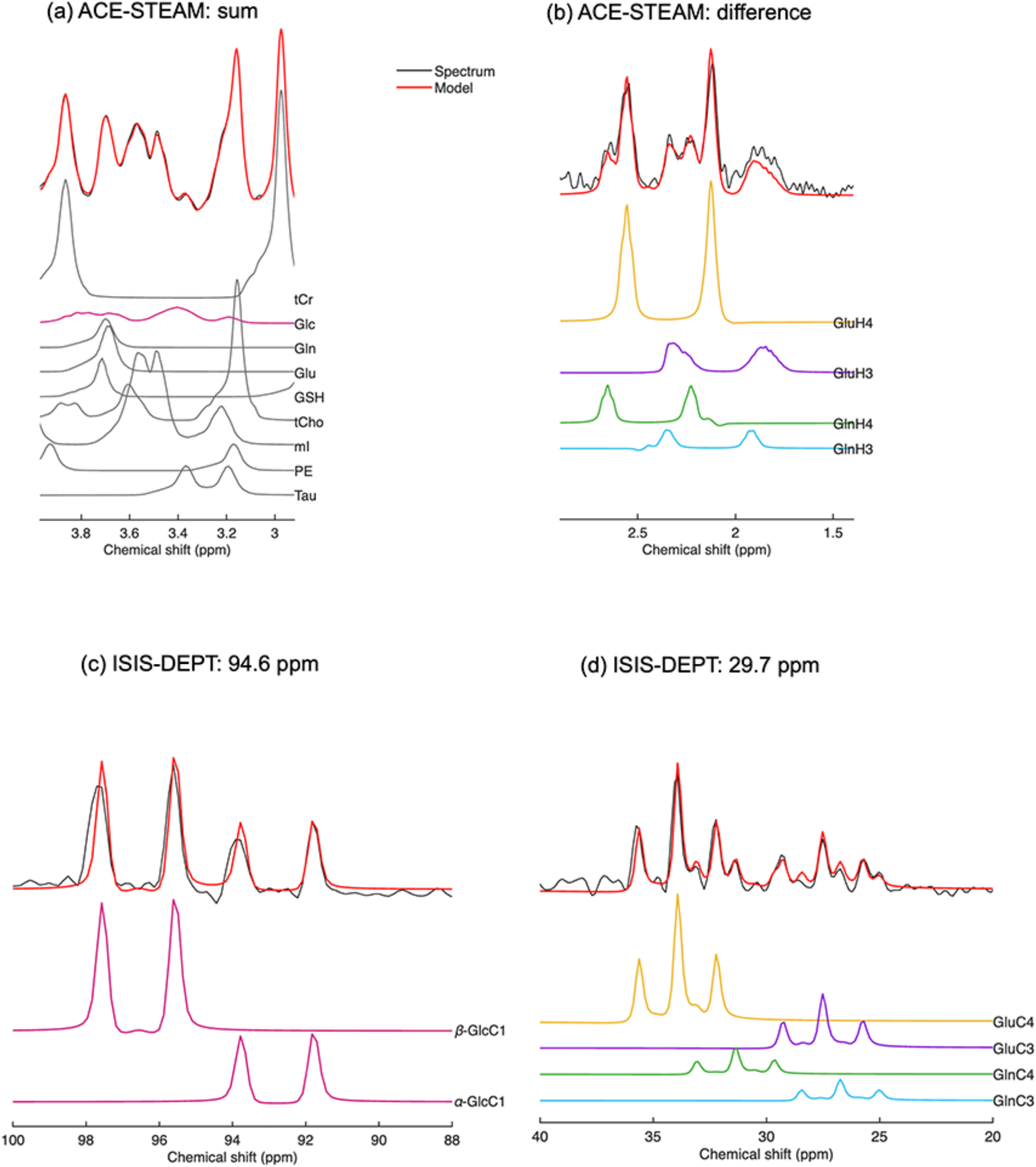
Representative in vivo spectra (black) acquired with ACE-STEAM and ISIS-DEPT sequences and sum resonances (red), as well as the individual isotopomer decompositions. (a) ACE-STEAM sum spectrum (32 averages). The spectrum shown here was processed with macromolecule and baseline removal, achieved by subtracting the fitted components obtained from the LCModel fitting results; (b) ACE-STEAM difference spectrum (128 averages); (c) ^13^C spectrum acquired with ISIS-DEPT (128 averages, carrier frequency 94.6 ppm); (d) ^13^C spectrum acquired with ISIS-DEPT (128 averages, carrier frequency 29.7 ppm).

At the end of infusion (60 min), the relative estimated fitting uncertainties (EFUs, from the estimation error bounds reported by LCModel) were ∼8% for β-GlcC1, ∼5% for GluH4, ∼29% for GlnH4, and ∼14% for GlxH3. The in vivo J_CH_ coupling constants derived from the ^13^C MR spectra were 146.0 Hz (GlcC1), 126.8 Hz (Glu/GlnC4), and 130.5 Hz (Glu/GlnC3), consistent across participants and similar to previous reports in the rat brain at 9.4 T^41^.

### 13C FEs and glucose concentration

As illustrated in Figure 4(a), despite the limited number of sampling points, the whole blood glucose levels of participants 2 and 3 display a characteristic rise followed by a decline over the course of the infusion and during the post-infusion period. Brain glucose reached a peak of 2.94 ± 0.04 mM at about 20-25 min after the start of the infusion, as shown in Figure 4(b), and then gradually declined toward baseline values by the end of the infusion (60 minutes). Blood samples were collected 4 times during the study for participants 2 and 3. In these two individuals, blood glucose rapidly increased from 6.2 ± 0.2 mM at baseline to 9.9 ± 2.2 mM within the first 15 min after bolus infusion, before declining to near baseline levels at ∼60 min. We omitted blood sampling for Participant 1 because the catheter clotted during the scan, and subsequent attempts at re-cannulation failed. However, the cerebral glucose response measured using MRS aligned with Participants 2 and 3, confirming consistency in brain glucose uptake across the participants under this protocol.

**Figure 4:**
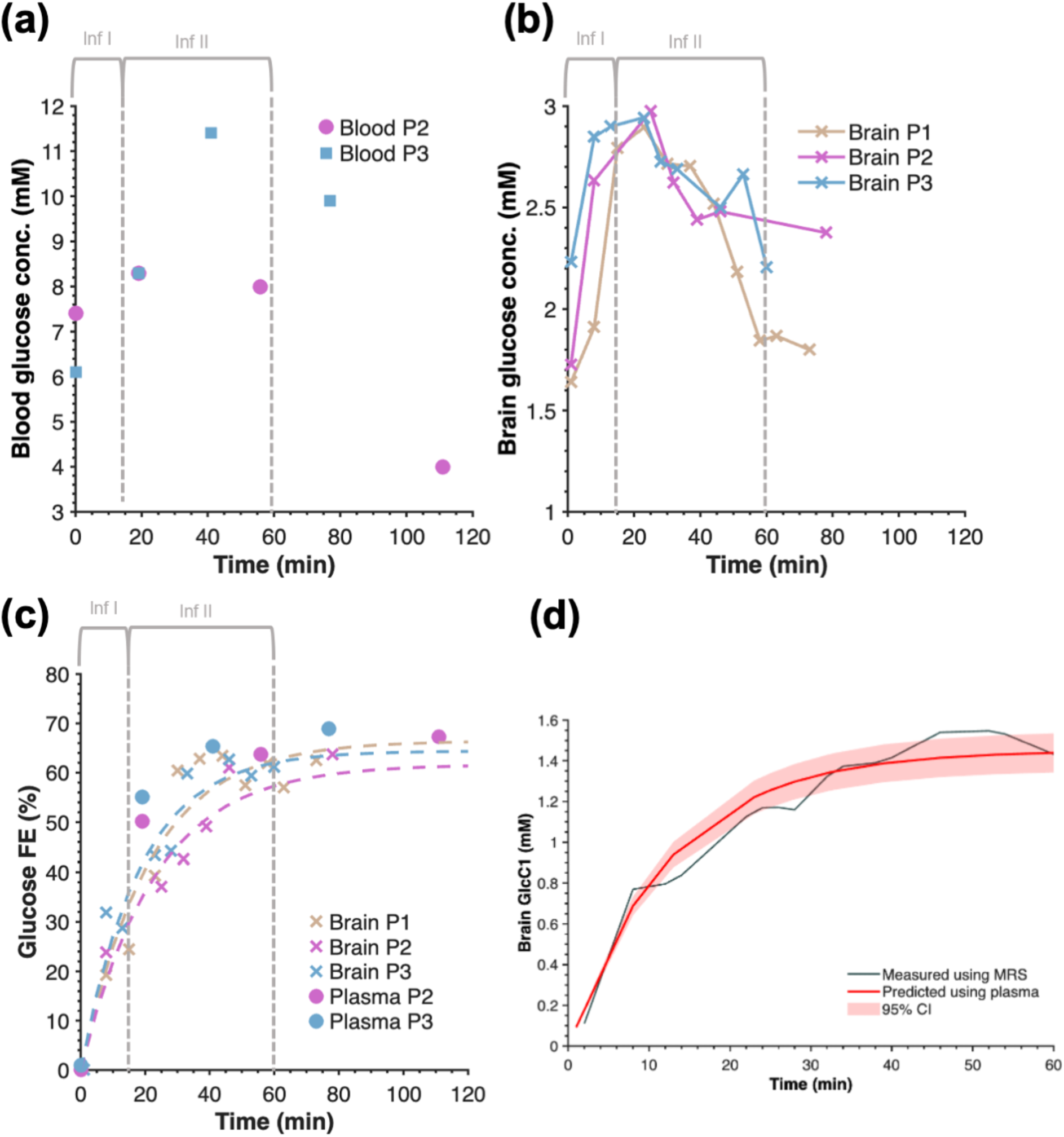
Time courses of glucose levels and ^13^C FEs. (a) blood glucose concentrations of Participant 2 and 3 measured with the glucose meter. (b) total brain glucose levels in three participants, quantified from ACE-STEAM sum spectra. (c) brain and plasma [1-^13^C]-glucose FEs measured using MRS and LC-HRMS, respectively. The dotted line represents the exponential fit of the brain data. (d) averaged brain [1-^13^C]-glucose (black) from Participant 2 and 3, and the fit of the model to the brain [1-^13^C]-glucose level using plasma data (red) with its 95% confidence interval (CI), using the reversible Michaelis-Menten model.

Across all three participants, this protocol yielded a reproducible glucose FE function for kinetic modeling (Table 1). Brain glucose FEs reached a maximum of 57-64% as shown in Figure 4(c) at the end of infusion. The mean rate constant (b = −0.05, time constant ∼20 min) indicates a rapid early rise in enrichment following the 14-min priming infusion. Plasma ^13^C glucose FEs, measured in Participants 2 and 3 (no plasma data were available for Participant 1), showed similar temporal dynamics but were slightly higher, reaching a steady state of 68.1 ± 1.1%. Fitting the reversible Michaelis-Menten model^42^ to the averaged FE time courses from Participants 2 and 3 yielded a relative glucose transport rate of T_max_/CMR_glc_ = 3.8 ± 0.2 with K_t_ = 0.6 mM, as shown in Figure 4(d).

**Table 1:** Individual and averaged V_TCA_ estimates and fitted GlcC1 FE parameters (a, b). V_TCA_ was computed for the group-average and individual, with uncertainties obtained from Monte-Carlo simulations, incorporating variability in GluC4, GlnC4, and GlxC3 enrichments. GlcC1 FE curves were fitted using the model GlcC1(t) = a(1 - exp(bt)).

| Participant | $V_{TCA}$ ($\mu\text{mol/g/min}$) | a | b ($\text{min}^{-1}$) |
| --- | --- | --- | --- |
| Average | $0.66 \pm 0.07$ | 0.64 | -0.05 |
| P1 | $0.58 \pm 0.05$ | 0.66 | -0.05 |
| P2 | $0.61 \pm 0.12$ | 0.62 | -0.04 |
| P3 | $0.73 \pm 0.10$ | 0.64 | -0.05 |

### TCA cycle flux estimation

By the end of the infusion, the mean FEs measured across the three participants were 0.179 ± 0.007 for GluC4, 0.140 ± 0.011 for GlnC4, and 0.076 ± 0.012 for GlxC3, reflecting progressive ^13^C incorporation into glutamate and glutamine pools.

Joint fitting of the average labeling of glutamate and glutamine yielded metabolic flux estimates of V_TCA_ = 0.66 ± 0.07 μmol/g/min, V_X_ = 0.88 ± 0.25 μmol/g/min, and V_GLN_ = 0.23 ± 0.08 μmol/g/min. The corresponding fitted curves for the average data are displayed in Figure 5. Participant-specific V_TCA_ values, obtained via Monte-Carlo simulations with V_X_ and V_GLN_ fixed to group means, ranged from 0.58 to 0.73 μmol/min/g (Table 1), and the individual fits of FE curves in the Supplementary material section 7. The measured V_TCA_ values fall within the range reported previously for the resting human cortex using various ^13^C MRS combined with blood sampling (typically 0.5-0.8 μmol/g/min, Supplementary material section 8).

**Figure 5:**
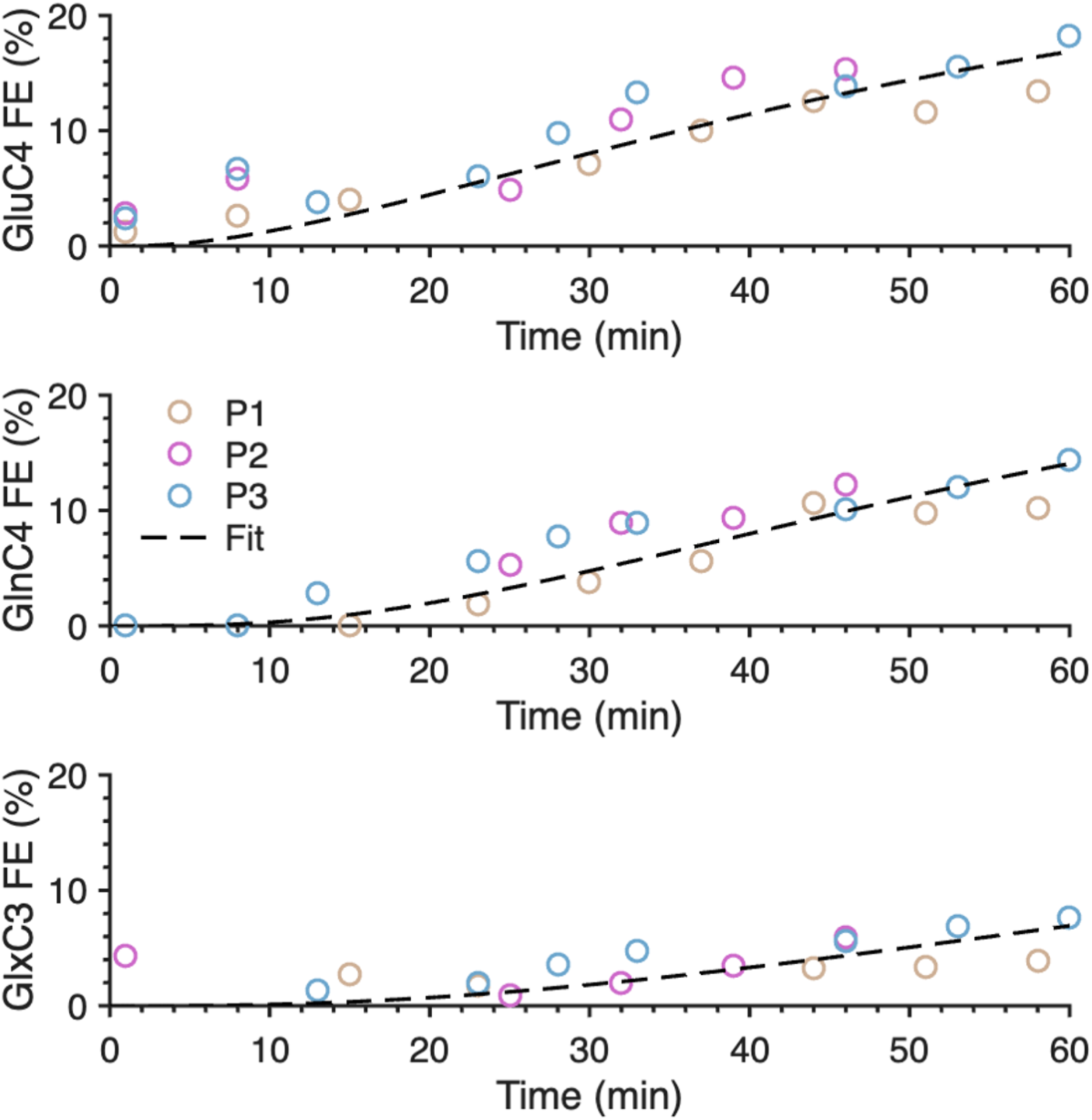
Average fits of glutamate C4, glutamine C4, and glutamate/glutamine C3 FE curves of 3 participants. The fitting was performed with the one-compartment model using the average GlcC1 function as input. Each participant is represented by a distinct color.

## Discussions

This study explored a clinically translatable protocol for dynamic, less-invasive quantification of energy metabolism in the human frontal lobe using interleaved ^1^H-[^13^C] and ^13^C-[^1^H] MRS at 7 T. This study demonstrated for the first time the time-resolved acquisition of both 1H and 13C MR spectra within a single experimental protocol. Through optimized MRS acquisition schemes, moderate-dose [1-^13^C]-glucose infusion, and a dual-tuned RF surface coil tailored for the frontal lobe, we successfully captured both cerebral glucose input and labeling curves of GluC4, GlnC4, GlxC3 with 5-7 min temporal resolution. The estimated VTCA values (0.58-0.73 μmol/g/min) align with most of the previous reports^12,14,15,16,17,18,22,28,48,49^ (Supplementary material section 8), and also comparable to the value obtained with ^2^H MRSI (0.60±0.13 μmol/g/min for the whole brain)^50^, confirming the feasibility of this approach for quantitative assessment of cerebral glucose oxidative metabolism.

### Blood sampling

In preclinical studies, simultaneous measurements of plasma and brain ^13^C glucose dynamics with high temporal resolution allow for estimation of both glucose transport and consumption rates.^45^ In humans, however, the conventional methods infer cerebral glucose FE from plasma using Michaelis-Menten kinetics, which assume rapid and saturated transport across the blood-brain barrier.^42^ Blood sampling can be challenging due to variable vein conditions and clotting. In this study, sampling failed for Participant 1 and required an extended time for the last two samples for Participant 2, disrupting scan timing and reducing MRS data points. To overcome this, we used direct in vivo measurement of brain GlcC1 as the input function. Its time course matched plasma-derived dynamics, supporting its reliability for modeling. This approach eliminates blood sampling, simplifies the protocol, and enables continuous MRS acquisition with improved temporal resolution.

### Glucose administration

Many of earlier ^13^C MRS studies used 20% (w/v) glucose solutions (∼1100 mOsm/kg)^17,43^, exceeding the generally accepted tolerance threshold (∼600 mOsm/kg) for peripheral infusion. To comply with safety guidelines, we employed a 10% (w/v) [1-^13^C]-glucose solution (∼620 mOsm/kg), within the acceptable clinical range^28^. This two-phase infusion strategy (initial bolus followed by slower maintenance) achieved stable cerebral enrichment while maintaining blood glucose within physiological limits^44^.

Moreover, the total administered glucose dose in our study was 0.5 g/kg body weight over 60 minutes, thereby lowering material costs. The initial priming infusion rapidly increased blood glucose to ∼10 mM, while the subsequent slower infusion maintained steady brain glucose FE and produced measurable labeling in glutamate and glutamine. ^13^C incorporation into GluC4, GlnC4, GluC3, and GlnC3 reached 7-17% over 1 hour, consistent with previous studies^13^.

Because brain [1-^13^C]-glucose reached a fractional enrichment of about 60-64%, the maximum achievable enrichment of [3-^13^C]-pyruvate would be approximately half of this value. It is therefore plausible that unlabeled substrates contribute substantially to cortical acetyl-CoA formation^45^, thereby diluting glutamate labeling to a maximum of ∼20%. Moreover, the ^13^C labeling of amino acids in this study did not reach isotopic steady state. In contrast, studies employing much higher [1-^13^C]-glucose infusion rates in the human cortex report the attainment of steady state after approximately 80 minutes of infusion.^16^ Our more moderate infusion protocol was chosen to improve participant safety and comfort while also reducing material costs.

### Isotopomer analysis

A strength of our approach is the absence of heteronuclear decoupling in any of the MRS sequences employed, offering the possibility for wide adoption in diverse clinical settings. Omission of decoupling to reduce RF power deposition and allowing faster repetition results in 1:1 doublets and 1:2:1 triplets from J_CH_ coupling in both ^1^H and ^13^C spectra. Despite this, all ^13^C-labeled signals were detectable within a two-minute acquisition after 30 minutes.

Unlike prior direct ^13^C MRS studies that relied on external phantoms for quantification^27^, our approach used the GluC4 as an internal normalization link between ISIS-DEPT and ACE-STEAM data. By jointly quantifying the last point GluC4 and GluH4, absolute ^13^C metabolite concentrations were derived from tCr-referenced ^1^H data, thereby canceling the dependence on external standards and reducing systematic errors that may be induced by non-uniform coil sensitivity across different loadings and inconsistent relaxation properties of metabolites across different chemical environments (e.g., water or tissues).

In principle, total glucose concentration and FE could be measured from the downfield α-Glc-H1 resonance (∼5.3 ppm), where one of the ^13^C-coupled peaks appears around 5.5 ppm. However, at 7T, reliable detection is challenging due to water residuals interfering with quantification, the low fractional population of α-Glc (∼36%), and attenuation from magnetization exchange, which reduces the ^13^C-coupled GlcH1 signal to near the MRS detection limit (∼0.1 mM). Therefore, total glucose was quantified from the 3-4 ppm region of ^1^H MR spectra, combined with GlcC1 measured using ISIS-DEPT, providing more robust and reproducible FE functions.

Although brain glucose was not strictly at steady-state during measurements, the model assumes constant flux through CMR_glc_. This is reasonable since glucose levels remained >1.6 mM, well above the hexokinase K_M_ (∼50 µM), where the enzyme operates near saturation and CMR_glc_ is relatively insensitive to glucose fluctuations. Consequently, downstream metabolic fluxes remain approximately constant.^45^ The T_max_/CMR_glc_ of 3.8 exceeded prior reports because blood because we measured glucose in blood and not plasma glucose (would be 10-15% larger), and we did not account for the contribution of blood glucose to the measured cerebral signal (would result in 15-20% less brain glucose).^46^ However, these factors do not affect V_TCA_ estimation.

We did not account for J_CC_ homonuclear coupling in our analysis of the ^13^C spectra. Although this could be considered a limitation, it should be noted that there is low ^13^C incorporation in our study using [1-^13^C]-glucose *versus* preclinical studies with [1,6-^13^C_2_]-glucose.^11,47^ Thus, while labeling rates in C4 are below 20%, the multiplicity arising from ^13^C homonuclear coupling is typically below the detection limit (within the level of spectral noise) in the absence of decoupling, suggesting that the impact on our measurements is likely negligible.^47^

Although the current study is limited to three participants, the results indicate that a 75-minute acquisition provides sufficient sensitivity for reliable estimation of metabolic fluxes. For method validation, we applied a one-compartment kinetic model to quantify TCA cycle. While Participants 1 and 2 underwent 3-sequence interleaved scans from the start, this was unnecessary due to low enrichment (<10%) in the first 30 minutes. Consequently, for Participant 3, early ISIS-DEPT at 29.7 ppm was omitted to prioritize input function measurements; deriving FE from ACE-STEAM difference spectra instead enhanced temporal resolution. Results confirmed that ISIS-DEPT can resolve GluC3 and GlnC3 after 30 minutes of infusion, which is not possible with indirect detection. Extending acquisitions beyond one hour will allow for distinct labeling curves, further refining flux determination in future models, whereas reducing scan time to better align with clinical study is possible depending on the required precision of flux estimates, but at the cost of increased uncertainty.

### Conclusion

This study demonstrates the feasibility and utility of a 7T interleaved ^1^H/^13^C MRS protocol, integrating an optimized RF coil and glucose administration protocol for quantifying cerebral glucose oxidative metabolism in the human frontal lobe. By enabling direct measurement of the brain glucose input function and ^13^C label incorporation into key metabolites, the method eliminates the need for blood sampling to access metabolic fluxes. This simplified approach offers a valuable platform for investigating metabolic dysregulation in neurological and psychiatric disorders.

## Data Availability

All data produced in the present study are available upon reasonable request to the authors.

## Acknowledgement

We acknowledge the MRI Platform of the Fondation Campus Biotech Geneva and the CIBM Center for Biomedical Imaging for providing expertise and resources to conduct this study. We greatly appreciate that Dr. Bernard Lanz provided us with his expertise throughout this project. We also acknowledge Dr. Tirone Fabiana and the Clinical Trial Unit of the Geneva University Hospital Pharmacy (la Pharmacie des HUG – Secteur des Essais Cliniques) for their assistance in the preparation of infusion materials. We are grateful to Dr. Laura Frangeul and the Human Cellular Neuroscience Platform (HCNP) for their support with blood processing. Special thanks to Dr. Julijana Ivanisevic and the Metabolomics and Lipidomics Platform of the Faculty of Biology and Medicine, University of Lausanne, for conducting the blood analyses. We would also like to express our gratitude to Yves Pilloud, Dr. Thanh Phong Lê, and Dr. Tan Toi Phan for their help on this project.

