## Supplementary Material for "Assessment of Glucose Metabolism *In Vivo* in the Human Frontal Lobe Using Interleaved ^1^H and ^13^C MRS at 7T: Toward Clinical Translation"

#### Section 1. Glucose infusion protocol

In this study, the first half glucose was infused to elevate plasma glucose concentration by 7 mM (126.8 mg/dL) above the individual's basal level within the first 14 minutes, and then the second half of glucose was used to slow the decline of glycemic level and preserve the brain glucose FE for the subsequent 44 minutes.

For example, for a participant weighing 53 kg, the priming dose was 13.2 g. The infusion plan of this phase followed the three-stage, variable-rate priming approach adapted from the protocol described by DeFronzo et al. (1979).

For all participants, the priming (I) and maintenance (II) doses were calculated individually and scaled to body weight, ensuring the total glucose administered was approximately 0.5 g/kg body weight. In this study, glucose was administered as a 10% (w/v) solution via a calibrated infusion pump.

Glucose infusion rate (GIR) can be calculated through the following equation: GIR (mg/kg/min) = glucose quantity (g) x 1000 / body weight (kg) / infusion time (min).

The table below presents an example of the complete infusion protocol over the whole 60-minute study, including the staged priming rates and constant maintenance rate, for a representative participant with a body weight of 53 kg:

| **Participant weight: 53 kg** | | | |
|---|---|---|---|
| **I-1** | glucose quantity | 7.20 | g |
| | volume | 72.00 | mL |
| | time | **4** | min |
| | GIR | **33.96** | mg/kg/min |
| | speed | **0.30** | mL/s |
| **I-2** | glucose quantity | 3.90 | g |
| | volume | 39.00 | mL |
| | time | **5** | min |
| | GIR | **14.72** | mg/kg/min |
| | speed | **0.13** | mL/s |

| | | | |
|---|---|---|---|
| **I-3** | glucose quantity | 2.10 | g |
| | volume | 21.00 | mL |
| | time | **5** | min |
| | GIR | **7.92** | mg/kg/min |
| | speed | **0.07** | mL/s |
| **II** | glucose quantity | 13.2 | g |
| | volume | 132 | mL |
| | time | **44** | min |
| | GIR | **5.66** | mg/kg/min |
| | speed | **0.05** | mL/s |
| **total** | glucose quantity | 26.40 | g |
| | volume | 264.00 | mL |
| | time | 58 | min |
| | total dosage | 0.50 | g/kg |

*Table S1.1. Detailed infusion plan over a 1-hour experiment for a representative participant with a body weight of 53 kg, outlining the timing, glucose infusion rates, and flow speed for each stage of the infusion.*

### Section 2. $^{1}H/^{13}C$ RF surface coil

#### Coil design

The 3/2-channel $^{1}H/^{13}C$ surface coil (Figure S2.1) consists of three 10 cm diameter circular $^{1}H$ loops and two 8.5 cm diameter circular $^{13}C$ loops made from the copper tubes. Both $^{1}H$ and $^{13}C$ coil arrays were implemented as transmit/receive (Tx/Rx) configurations without additional transmit-only loops. $^{13}C$ excitation was performed in quadrature mode using two channels, while the $^{1}H$ array was driven using a fixed three-channel transmit configuration via RF power splitting. Although the RF interface (Stark Contrast, Erlangen, Germany) supports additional inter-nuclear decoupling functions, these were not employed in this study, and channel coupling was determined solely by the intrinsic coil design. The mutual coupling between the loops was minimized by overlapping.

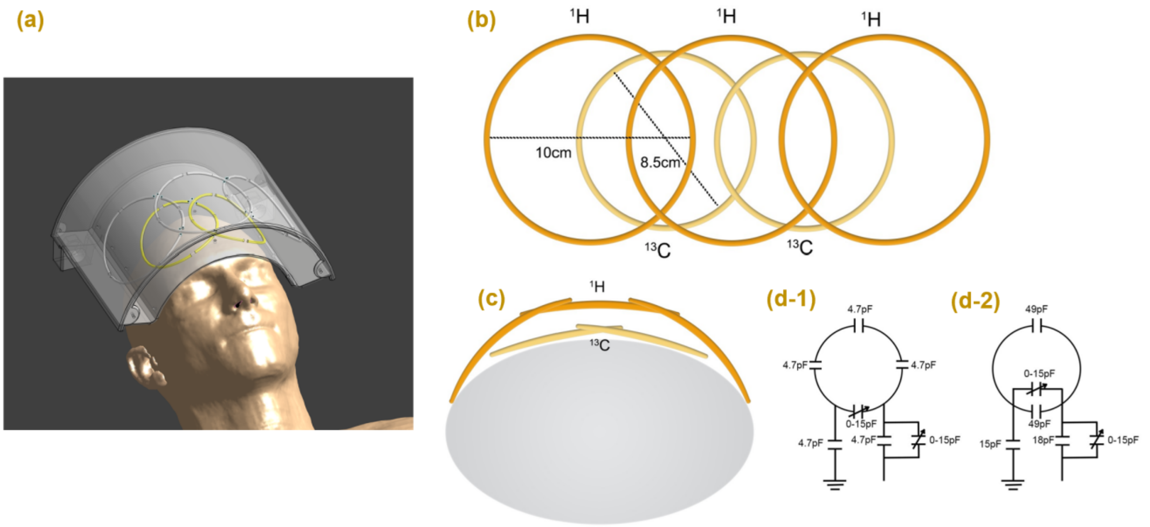


*Figure S2.1. Configuration of the 3-channel $^{1}H$/2-channel $^{13}C$ RF coil. (a) All loops are enclosed within the casing to cover the human frontal lobe (screenshot in Sim4Life); (b) schematic top view: the $^{1}H$ loops with a diameter of 10 cm above the $^{13}C$ loops with a diameter of 8.5 cm; (c) schematic side view: geometrical isolation between $^{13}C$ and $^{1}H$ loops; (d-1) Design and capacitor values for $^{1}H$ loop; (d-2) Design and capacitor values for $^{13}C$ loop.*

Electromagnetic simulations were performed using the finite-difference time-domain (FDTD) solver in Sim4Life (ZMT Zurich MedTech, Switzerland) for both the $^{1}H$ and $^{13}C$ RF coils under realistic loading conditions using the Duke human voxel model (IT'IS Virtual Population, Christ et al., 2010). For each coil, SAR analysis was conducted for the prescribed transmit configuration (quadrature mode for $^{13}C$ and fixed 3-channel transmit mode for $^{1}H$). Local SAR (10 g averaged) and whole-body average SAR were computed and normalized to the unit accepted power and subsequently scaled to the operating transmit conditions. Compliance with IEC 60601-2-33 guidelines was verified by ensuring SAR limits for Normal Operating Mode were not exceeded. A conservative transmit power calibration was applied using simulation-derived k-factors without explicit correction for full RF chain losses, together with additional safety margins (Steensma et al., 2023) accounting for inter-subject variability (factor 1.4) and directional coupler calibration uncertainty (10%).

#### Bench measurement

Bench measurements (Figure S2.2) were performed on four volunteers to evaluate the impact of variable subject loading on coil performance. S-parameters, specifically $S_{11}$ (reflection) and $S_{21}$ (transmission), were measured with volunteers positioned supine inside the RF shielding. Both $^{13}$C channels showed consistent matching across all subjects, with $S_{11}$ values ranging from –19.6 to –24.3 dB, indicating low reflection and good impedance matching. All inter-channel isolation ($S_{21}$) remained about –20 to -30 dB across all subjects, confirming effective decoupling between different coil elements. Minor frequency shifts of < 0.3 MHz were observed with different subject loading, which indicates the coil did not require retuning at the time of use.

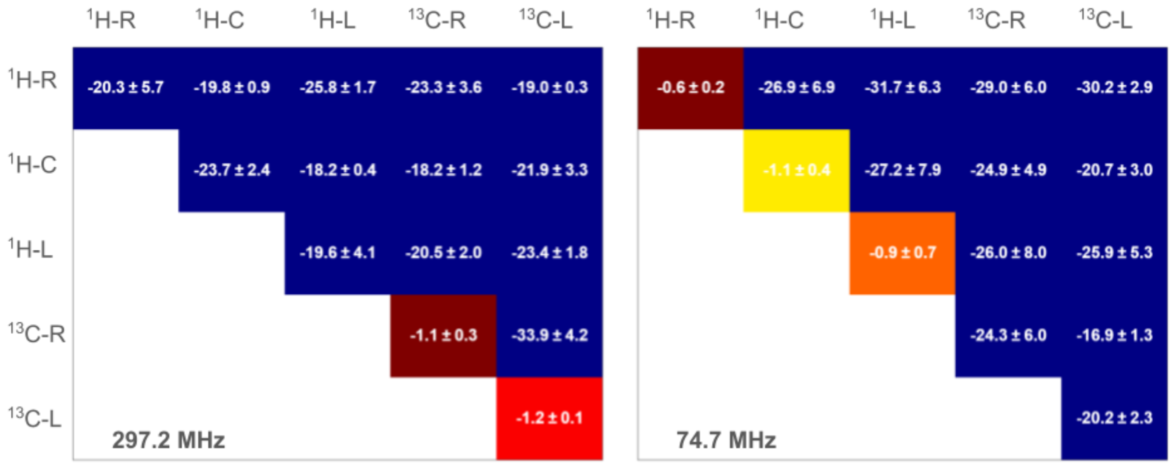


*Figure S2.2. S-parameters measured from a network analyzer at 7T. Each cell represents the mean ± standard deviation of S-parameters (in dB) measured across four volunteers using the dual-tuned $^{1}H/^{13}C$ RF coil. Measurements were performed at the Larmor frequencies of $^{1}$H (297.2 MHz, left) and $^{13}$C (74.7 MHz, right).*

### Section 3. Sequence design

#### ACE-STEAM (Adiabatic Carbon-Edited single-voxel Stimulated Echo Acquisition Mode)

The ACE-STEAM sequence (shown in Figure S3.1) was adapted from the ACED-STEAM protocol described by Pfeuffer et al. (1999). Water suppression (WS) was achieved using a variable pulse power and optimized relaxation delay (VAPOR) scheme (Tkáč et al., 1999), consisting of eight Gaussian RF pulses with flip angles of 90.0°, 90.0°, 160.2°, 160.2°, 143.1°, 90.0°, 160.2°, and 160.2°. The corresponding inter-pulse delays were 150, 100, 122, 105, 102, 61, 69, and 14 ms. Three blocks of outer-volume saturation (OVS) were inserted into the gaps between the last four water-suppression pulses, following a *y–y–xyz* scheme. All OVS pulses were adiabatic hyperbolic secant pulses with a 90° flip angle.

The slice-selective RF pulse was designed using the Shinnar–Le Roux (SLR) algorithm (Balchandani et al., 2010). The asymmetric, amplitude-modulated pulse had a duration of 1.6 ms, a bandwidth of 2 kHz at $\gamma B_1/2\pi$ of 1.0 kHz. An additional water-suppression pulse (15-ms Gaussian, flip angle = 160.8°) was applied after the second excitation pulse in the STEAM sequence.

TE was set to 7.9 ms, corresponding to $1/J_{CH}$, to achieve the spectral resonance editing. The mixing time (TM) was fixed at 35 ms. The $^{13}C$ 180° inversion pulse was implemented as an $HS_4$ adiabatic pulse (duration: 6.656 ms, carrier frequency at 29.7 ppm), as shown below. This editing pulse was alternately toggled on and off, enabling difference-spectrum acquisition.

All crusher gradients were optimized using the DOTCOPS algorithm (Landheer et al., 2019) to minimize residual transverse magnetization. A 32-step phase-cycling scheme was employed to suppress unwanted coherence pathways. The specific phase-cycling matrix is shown below.

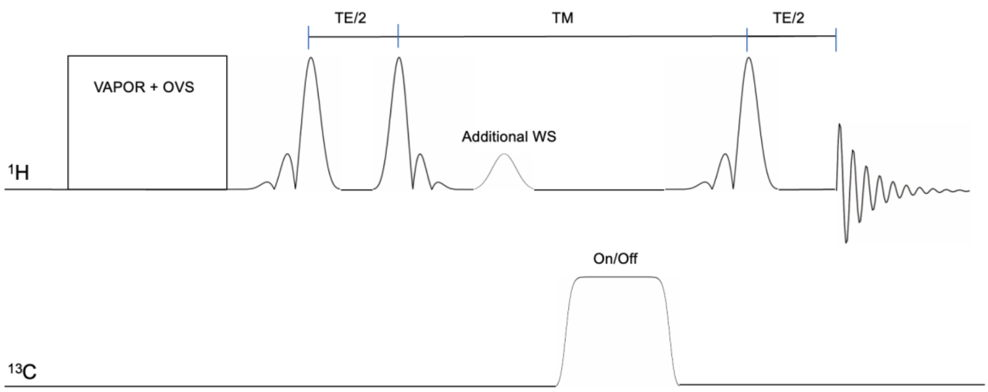


*Figure S3.1. ACE-STEAM pulse sequence diagram.*

#### ISIS-DEPT (Image-Selected In Vivo Spectroscopy combined with Distortionless Enhancement by Polarization Transfer)

The ISIS-DEPT sequence was implemented as illustrated in Figure S3.2. The OVS module was implemented using the same approach as in the ACE-STEAM sequence. For $^{1}H$ localization, three identical $HS_4$ pulses along respectively on x, y, and z, was employed, providing an inversion bandwidth of 10.06 kHz at a $\gamma B_1/2\pi$ of 1.0 kHz. Proton polarization transfer to $^{13}C$ was achieved using 200 μs rectangular 90° pulses for $^{1}H$ excitation. Following the $^{1}H$ 180° rectangular inversion pulse (400 μs), a 100 μs delay was introduced before applying the $^{13}C$ pulses.

For $B_1$-insensitive rotation, the segmented 0° BIR-4 adiabatic pulses (de Graaf et al., 1995, 6.4-ms duration, bandwidth = 1.25 kHz at $\gamma B_1/2\pi$ = 1.0 kHz) were applied on the $^{13}C$ frequency. An inter-segment delay of 2.1 ms was experimentally optimized so that the midpoint of the pulse occurred 3.7 ms after. The flip angle of the final $^{1}H$ θ pulse was adjusted based on the targeted carbon multiplicity: 90° for optimal enhancement of CH groups (for [1-$^{13}C$] glucose detection) and 45° for $CH_2$ groups (for [3-$^{13}C$] glutamate, [4-$^{13}C$] glutamate, [3-$^{13}C$] glutamine, [4-$^{13}C$] glutamine detection).

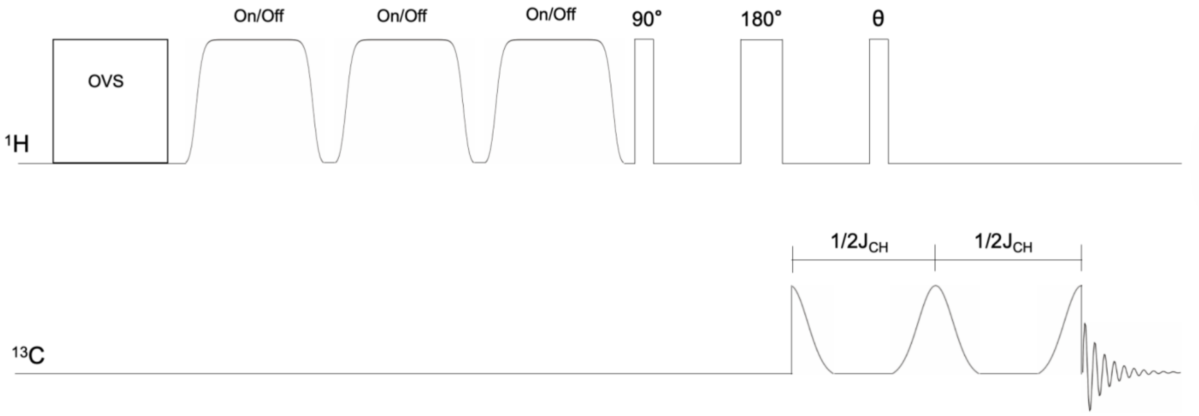


*Figure S3.2. ISIS-DEPT pulse sequence diagram.*

### Section 4. MRSinMRS checklist

| 1. Hardware | | |
|---|---|---|
| a. Field strength [T] | 7 T | 7 T |
| b. Manufacturer | Siemens | Siemens |
| c. Model (software version if available) | XA60 | XA60 |
| d. RF coils: nuclei (transmit/ receive), number of channels, type, body part | 3/2-channel $^{1}H/^{13}C$, surface coil, head | 3/2-channel $^{1}H/^{13}C$, surface coil, head |
| e. Additional hardware | N/A | N/A |
| **2. Acquisition** | | |
| a. Pulse sequence | ACE-STEAM | ISIS-DEPT |
| b. Volume of Interest (VOI) locations | Frontal lobe | Frontal lobe |
| c. Nominal VOI size [cm$^3$, mm$^3$] | 2.8 × 2.5 × 4.0 cm$^3$ | 3.0 × 3.0 × 4.0 cm$^3$ |
| d. Repetition Time (TR), Echo Time (TE) [ms, s] | TR = 4 s, TE = 7.9 ms | TR = 4 s |
| e. Total number of Excitations or acquisitions per spectrum<br><br>In time series for kinetic studies<br><br>i. Number of Averaged spectra (NA) per time-point<br>ii. Averaging method (e.g. block-wise or moving average)<br>iii. Total number of spectra (acquired / in time-series) | 32 averages per time-point, block-wise, 9-11 blocks | 32 averages per time-point, block-wise, 4 blocks |
| f. Additional sequence parameters<br><br>(spectral width in Hz, number of spectral points, frequency offsets)<br><br>If STEAM:, Mixing Time (TM)<br><br>If MRSI: 2D or 3D, FOV in all directions, matrix size, acceleration factors, sampling method | SW = 8000 Hz, 4096 pts, 2.3 ppm<br><br>TM = 35 ms | SW = 20000 Hz, 4096 pts, 29.7 and 94.6 ppm |
| g. Water Suppression Method | VAPOR | N/A |

| | | |
|---|---|---|
| h. Shimming Method, reference peak, and thresholds for "acceptance of shim" chosen | Vendor shimming method ("GRE-brain") and FAST(EST)MAP | Vendor shimming method ("GRE-brain") and FAST(EST)MAP |
| i. Triggering or motion correction method<br><br>(respiratory, peripheral, cardiac triggering, incl. device used and delays) | N/A | N/A |
| 3. Data analysis methods and outputs | | |
| a. Analysis software | MATLAB | MATLAB |
| b. Processing steps deviating from quoted reference or product | S/N$^2$ coil combination, spectral alignment to NAA, HSVD water suppression, truncated to 1024 points, zero-filling to 4096 points | S/N$^2$ coil combination, truncated to 1024 points, zero-filling to 4096 points, MP-PCA denoising, 5 Hz exponential line-broadening, phase correction, and frequency alignment to GlcC1 or GluC4 |
| c. Output measure<br><br>(e.g. absolute concentration, institutional units, ratio)Processing steps deviating from quoted reference or product | Ratio to tCr | Ratio to endpoint GluC4 |
| d. Quantification references and assumptions, fitting model assumptions | Assume baseline tCr concentration to be 8 mM. | Total GlcC1 was derived from the β-anomer, assuming a 64% abundance. The absolute concentration of β-GlcC1 was determined by scaling against the GluH4 concentration measured at the final time point using ACE-STEAM difference spectra and GluC4 from ISIS-DEPT (29.7 ppm). |
| 4. Data Quality | | |
| a. Reported variables<br><br>(SNR, Linewidth (with reference peaks)) | Water linewidth described | Chemical shifts of all peaks described |
| b. Data exclusion criteria | No data excluded | No data excluded |
| c. Quality measures of postprocessing Model fitting | The relative EFUs at 60 min described | The relative EFUs at 60 min described |

| | | |
|---|---|---|
| (e.g. CRLB, goodness of fit, SD of residual) | | |
| d. Sample Spectrum | Figure 2 (a) and (b) | Figure 2 (c) and (d) |

#### Section 5. Spin parameters

Chemical shifts and J-coupling constants for α-GlcC1, β-GlcC1, GluC4, GlnC4, GluC3, and GlnC3 spin systems. Values in parentheses indicate resonance chemical shifts in ppm. J-coupling constants are reported in Hz. The $^{1}H$ and $^{13}C$ chemical shifts and $J_{HH}$ coupling constants were referred to Govindaraju et al. (2000) and Henry et al. (2003). The values with * are the measurements of our study. A value of 0 indicates no coupling constant specified in the simulation.

##### α-GlcC1

| | H1 (5.230) | H2 (3.519) | H3 (3.689) | H4 (3.395) | H5 (3.822) | H6a (3.826) | H6b (3.749) | C1 (92.830) |
|---|---|---|---|---|---|---|---|---|
| H1 | 0.000 | 3.800 | 0.000 | 0.000 | 0.000 | 0.000 | 0.000 | 146.000* |
| H2 | 3.800 | 0.000 | 9.600 | 0.000 | 0.000 | 0.000 | 0.000 | 0.000 |
| H3 | 0.000 | 9.600 | 0.000 | 9.400 | 0.000 | 0.000 | 0.000 | 0.000 |
| H4 | 0.000 | 0.000 | 9.400 | 0.000 | 9.900 | 0.000 | 0.000 | 0.000 |
| H5 | 0.000 | 0.000 | 0.000 | 9.900 | 0.000 | 1.500 | 6.000 | 0.000 |
| H6a | 0.000 | 0.000 | 0.000 | 0.000 | 1.500 | 0.000 | −12.100 | 0.000 |
| H6b | 0.000 | 0.000 | 0.000 | 0.000 | 6.000 | −12.100 | 0.000 | 0.000 |
| C1 | 146.000* | 0.000 | 0.000 | 0.000 | 0.000 | 0.000 | 0.000 | 0.000 |

##### β-GlcC1

| | H1 (4.630) | H2 (3.230) | H3 (3.473) | H4 (3.387) | H5 (3.450) | H6a (3.882) | H6b (3.707) | C1 (96.62) |
|---|---|---|---|---|---|---|---|---|
| H1 | 0.000 | 8.000 | 0.000 | 0.000 | 0.000 | 0.000 | 0.000 | 146.000* |
| H2 | 8.000 | 0.000 | 9.100 | 0.000 | 0.000 | 0.000 | 0.000 | 0.000 |
| H3 | 0.000 | 9.100 | 0.000 | 9.400 | 0.000 | 0.000 | 0.000 | 0.000 |
| H4 | 0.000 | 0.000 | 9.400 | 0.000 | 8.900 | 0.000 | 0.000 | 0.000 |
| H5 | 0.000 | 0.000 | 0.000 | 8.900 | 0.000 | 1.600 | 5.400 | 0.000 |
| H6a | 0.000 | 0.000 | 0.000 | 0.000 | 1.600 | 0.000 | −12.300 | 0.000 |
| H6b | 0.000 | 0.000 | 0.000 | 0.000 | 5.400 | −12.300 | 0.000 | 0.000 |

| | | | | | | | |
|---|---|---|---|---|---|---|---|
| C1 | 146.000* | 0.000 | 0.000 | 0.000 | 0.000 | 0.000 | 0.000 | 0.000 |

#### GluC4

| | H1 (3.743) | H2 (2.037) | H3 (2.120) | H4 (2.337) | H5 (2.352) | C (34.000) |
|---|---|---|---|---|---|---|
| H1 | 0.000 | 7.331 | 4.651 | 0.000 | 0.000 | 4.520 |
| H2 | 7.331 | 0.000 | −14.849 | 6.413 | 8.406 | 4.310 |
| H3 | 4.651 | −14.849 | 0.000 | 8.478 | 6.875 | 4.310 |
| H4 | 0.000 | 6.413 | 8.478 | 0.000 | −15.915 | 126.790* |
| H5 | 0.000 | 8.406 | 6.875 | −15.915 | 0.000 | 126.790* |
| C | 4.520 | 4.310 | 4.310 | 126.790* | 126.790* | 0.000 |

#### GlnC4

| | H1 (3.753) | H2 (2.129) | H3 (2.109) | H4 (2.432) | H5 (2.454) | C (31.450) |
|---|---|---|---|---|---|---|
| H1 | 0.000 | 5.847 | 6.500 | 0.000 | 0.000 | 4.470 |
| H2 | 5.847 | 0.000 | −14.504 | 9.165 | 6.437 | 4.470 |
| H3 | 6.500 | −14.504 | 0.000 | 6.324 | 9.209 | 4.470 |
| H4 | 0.000 | 9.165 | 6.324 | 0.000 | −15.371 | 126.790* |
| H5 | 0.000 | 6.437 | 9.209 | −15.371 | 0.000 | 126.790* |
| C | 4.470 | 4.470 | 4.470 | 126.790* | 126.790* | 0.000 |

#### GluC3

| | H1 (3.743) | H2 (2.037) | H3 (2.120) | H4 (2.337) | H5 (2.352) | C (27.600) |
|---|---|---|---|---|---|---|
| H1 | 0.000 | 7.331 | 4.651 | 0.000 | 0.000 | 4.290 |
| H2 | 7.331 | 0.000 | −14.849 | 6.413 | 8.406 | 130.450* |
| H3 | 4.651 | −14.849 | 0.000 | 8.478 | 6.875 | 130.450* |
| H4 | 0.000 | 6.413 | 8.478 | 0.000 | −15.915 | 4.290 |

| | | | | | | |
|---|---|---|---|---|---|---|
| H5 | 0.000 | 8.406 | 6.875 | −15.915 | 0.000 | 4.290 |
| C | 4.290 | 130.450* | 130.450* | 4.290 | 4.290 | 0.000 |

#### GlnC3

| | H1 (3.753) | H2 (2.129) | H3 (2.109) | H4 (2.432) | H5 (2.454) | C (27.130) |
|---|---|---|---|---|---|---|
| H1 | 0.000 | 5.847 | 6.500 | 0.000 | 0.000 | 4.640 |
| H2 | 5.847 | 0.000 | −14.504 | 9.165 | 6.437 | 127.050* |
| H3 | 6.500 | −14.504 | 0.000 | 6.324 | 9.209 | 127.050* |
| H4 | 0.000 | 9.165 | 6.324 | 0.000 | −15.371 | 4.820 |
| H5 | 0.000 | 6.437 | 9.209 | −15.371 | 0.000 | 4.820 |
| C | 4.640 | 127.050* | 127.050* | 4.820 | 4.820 | 0.000 |

* Values measured in this study

#### Section 6. Mathematical metabolic model

To quantify the underlying metabolic fluxes, we utilized a one-compartment kinetic model adapted from literature (Shulman et al., 1995; Henry et al., 2005; Shen et al., 2014; and Xin et al., 2015). This model mathematically describes the time course of the $^{13}$C label as it moves from the glucose tracer into key metabolic intermediates and neurotransmitters.

By employing a system of ordinary differential equations ODEs, the model solves for the metabolic flux rates that best fit the measured data.

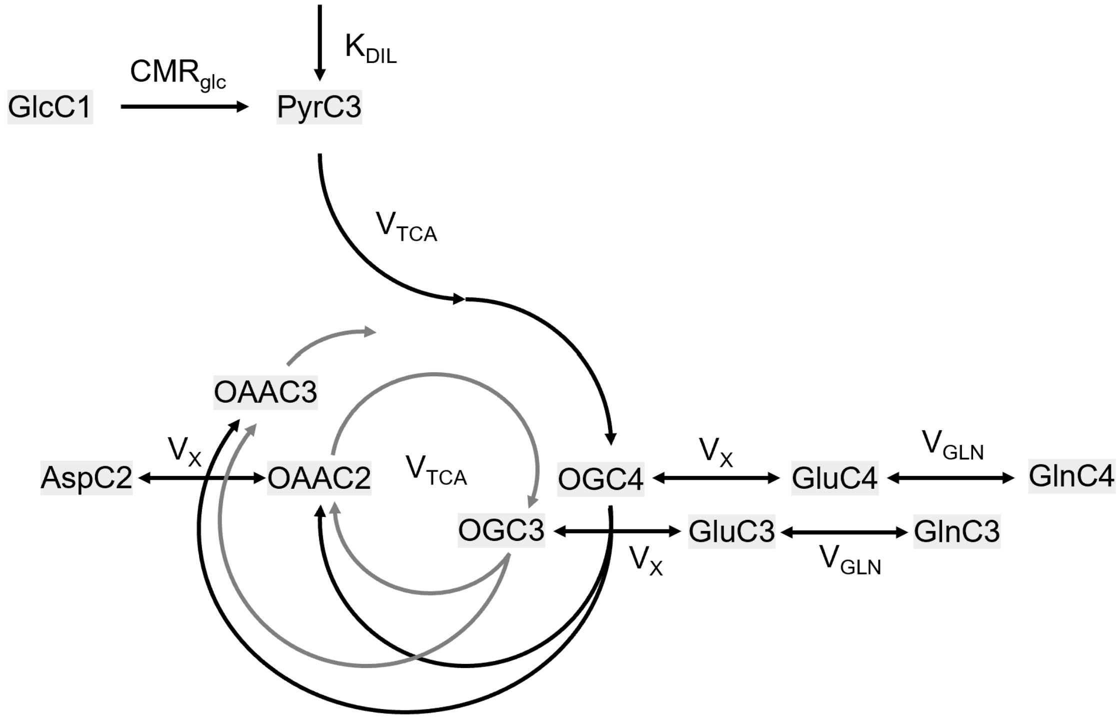


*Figure S5.1. Schematic of $^{13}$C fluxes following [1-$^{13}$C] glucose infusion with the one-compartment metabolic model. GlcC1, [1-$^{13}$C] glucose; PyrC3, [3-$^{13}$C] pyruvate; OGC4, [4-$^{13}$C] 2-oxoglutarate; OGC3, [3-$^{13}$C] 2-oxoglutarate; OAAC2, [2-$^{13}$C] oxaloacetate; OAAC3, [3-$^{13}$C] oxaloacetate; GluC4, [4-$^{13}$C] glutamate; GluC3, [3-$^{13}$C] glutamate; GlnC4, [4-$^{13}$C] glutamine; GlnC3, [3-$^{13}$C] glutamine; AspC3, [3-$^{13}$C] aspartate; $CMR_{glc}$, cerebral metabolic rate of glucose consumption; $K_{DIL}$, the dilution factor of the pyruvate pool induced by uptake of unlabeled substrate from plasma; $V_{TCA}$, TCA cycle rate; $V_X$, exchange flux between mitochondrial TCA cycle intermediates and cytosolic glutamate; $V_{GLN}$, glutamate-glutamine exchange rate.*

As shown in Figure S5.1, the metabolic fate of GlcC1 can be tracked through glycolysis and the TCA cycle. During glycolysis, GlcC1 is metabolized to PyrC3 at a rate corresponding to the cerebral metabolic rate of glucose ($CMR_{glc}$). However, initiating the TCA cycle requires accounting for dilution of labeled acetyl-CoA in the brain arising from the conversion of unlabeled substrates, including unlabeled pyruvate transported from the plasma, acetate and fatty acids metabolized within astrocytes. To account for this effect, a factor $K_{DIL}$ is introduced, which describes the dilution effects.

Following the conversion of pyruvate to acetyl-CoA, the $^{13}$C label enters the TCA cycle and is incorporated into intermediates such as OG. OG undergoes transamination and exchanges with the cytosolic glutamate pool, which in turn can interconvert with glutamine. During the first turn of the TCA cycle, the $^{13}$C label appears at OGC4 at a rate of $V_{TCA}$. OGC4 subsequently exchanges with GluC4 via mitochondrial transport with an exchange rate $V_X$. In the second TCA cycle turn, the label is transferred from OGC4 to OGC3 through OAAC2, resulting in further labeling of GluC3 and GlnC3 at a rate of $0.5V_{TCA}$, as half of the OGC4 label is lost to $OAA_3$. The glutamate–glutamine exchange proceeds with a rate constant $V_{GLN}$. And aspartate is labeled with the rate of $V_X$ as AspC2 via the transmitochondrial exchange with OAAC2.

The dynamic labeling of metabolites was modeled using the described one-compartment system comprising nine $^{13}$C-labeled metabolite concentrations (PyrC3, OGC4, OGC3, GluC4, GluC3, GlnC4, GlnC3, OAAC2, AspC2).

The free parameters estimated in the fitting procedure included:

- $V_{TCA}$
- $V_X$
- $V_{GLN}$

Additional fixed parameters for the fitting in this study were:

- $K_{DIL}$= 0.82, taken from Xin et al. (2015), and Lebon et al. (2002).
- $CMR_{glc}$: set to $0.5V_{TCA}$ based on pyruvate mass balance.

All metabolite pool sizes (PYR, OG, GLU, GLN, OAA, ASP) used in the ODE system remain constants. $FE_{GlcC1}$ represents the time-dependent fractional enrichment of labelled glucose measured in the brain. The free flux parameters were optimized by fitting the simulated $^{13}$C label time courses to the measured FE dynamics of GluC4, GlnC4, and GlxC3 using nonlinear least-squares estimation.

The ODE system:

PyrC3:

$$dPyrC3/dt = CMR_{glc} \times FE_{GlcC1}(t) - V_{TCA} \times PyrC3 / PYR$$

OGC4:

$$dOGC4/dt = V_{TCA} \times K_{DIL} \times PyrC3 / PYR + V_X \times GluC4 / GLU - (V_{TCA} + V_X) \times OGC4 / OG$$

OGC3:

$$dOGC3/dt = V_{TCA} \times OAAC2 / OAA - (V_{TCA} + V_X) \times OGC3/ OG + V_X \times GluC3 / GLU$$

GluC4:

$$dGluC4/dt = V_X \times OGC4 / OG - (V_X + V_{GLN}) \times GluC4 / GLU + V_{GLN} \times GlnC4 / GLN$$

GluC3:

$$dGluC3/dt = V_X \times OGC3 / OG - (V_X + V_{GLN}) \times GluC3 / GLU + V_{GLN} \times GlnC3 / GLN$$

GlnC4:

$$dGlnC4/dt = V_{GLN} \times (GluC4 / GLU - GlnC4 / GLN)$$

GlnC3:

$$dGlnC3/dt = V_{GLN} \times (GluC3 / GLU - GlnC3 / GLN)$$

OAAC2:

$$dOAAC2/dt = 0.5 \times V_{TCA} \times (OGC4 + OGC3) / OG + V_X \times AspC2 / ASP - (V_{TCA} + V_X) \times OAAC2 / OAA$$

AspC2:

$$dAspC2/dt = V_X \times (OAAC2 / OAA - AspC2 / ASP)$$

#### Section 7. Individual fits of FE curves

Individual fits of glutamate C4, glutamine C4, and glutamate/glutamine C3 FE curves of 3 participants. The fitting was performed with the one-compartment model using the individual GlcC1 function as input, with $V_X$ (=0.88 µmol/g/min) and $V_{GLN}$ (=0.23 µmol/g/min) fixed to the group-mean values.

P1:

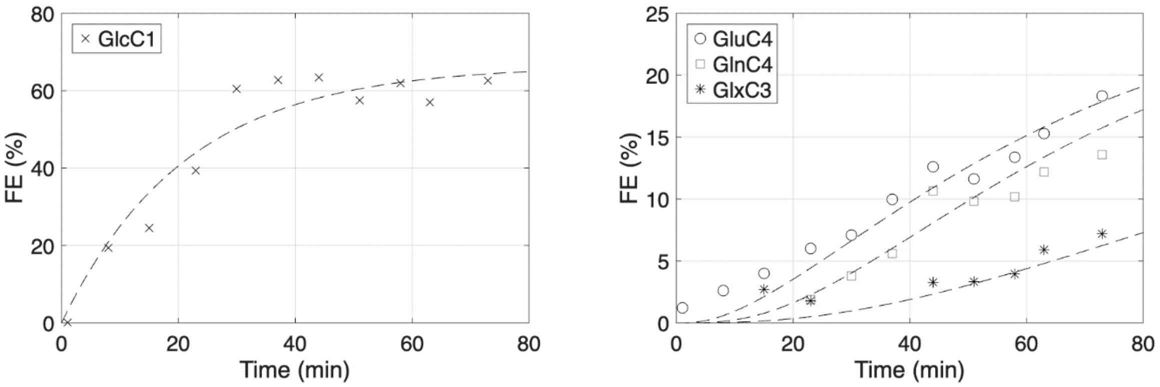


P2:

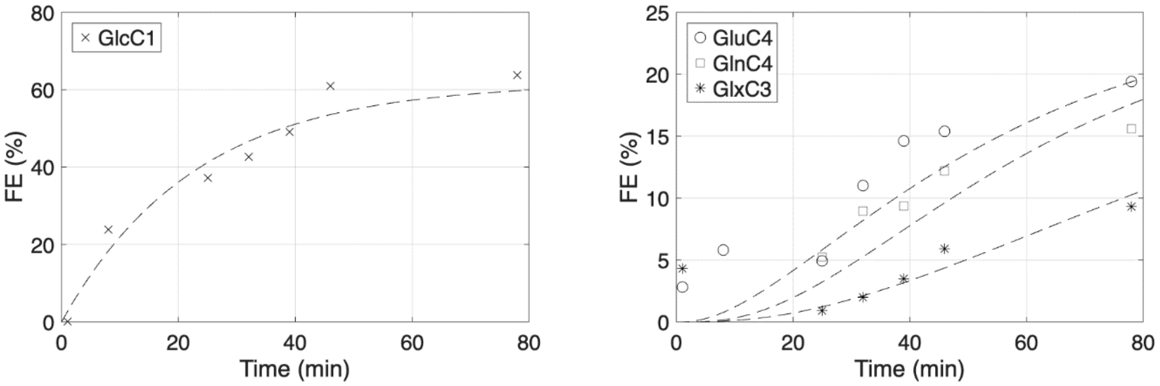


P3:

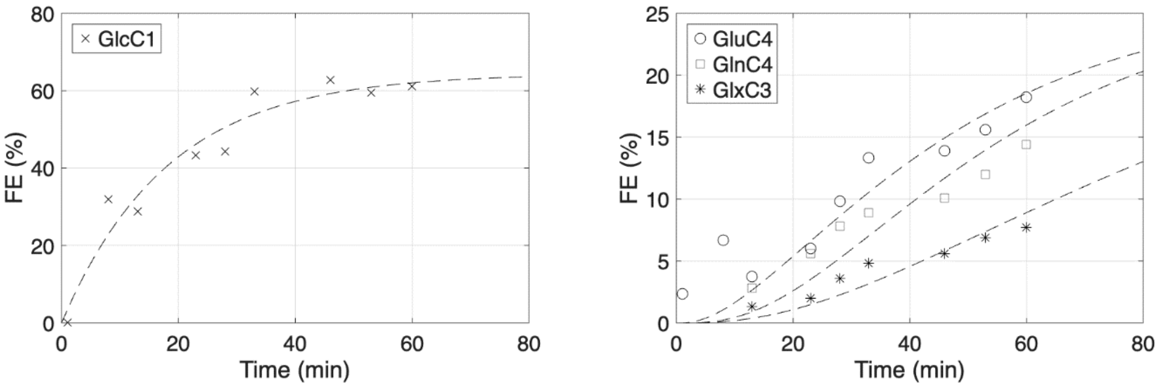

#### Section 8. TCA cycle flux in healthy human brains

TCA cycle flux in healthy human brains at resting state that were measured by $^{13}C$ (direct) or $^{1}H$-[$^{13}C$] MR spectroscopy (indirect).

| **Study** | **Brain region** | **$V_{TCA}$ (μmol/g/min)** | **Field strength, method** |
|---|---|---|---|
| Mason et al., 1995 | neocortex | 0.73 ± 0.19 | 2.1 T, direct |
| Shen et al., 1999 | occipital and parietal lobes | 0.77 ± 0.07 | 2.1 T, direct |
| Pan et al., 2000 | occipital lobe | 0.66 ± 0.09 | 4 T, indirect |
| Chhina et al., 2001 | visual cortex | 0.60 ± 0.10 | 3 T, direct |
| Chen et al., 2001 | visual cortex | 0.83 ± 0.13 | 4 T, indirect |
| Gruetter et al., 2001 | occipital lobe | 0.57 ± 0.06 | 4 T, direct |
| Mason et al., 2003 | occipital lobe | 0.52 ± 0.12 | 2.1 T, direct |
| Boumezbeur et al., 2010 | occipito-parietal lobe | 0.65 ± 0.03 | 4 T, direct and indirect |
| Dehghani et al., 2020 | anterior/posterior cingulate cortex | 0.23-0.29 (ACC)<br>0.85-1.18 (PCC) | 3 T, indirect |
| Ziegs et al., 2022 | occipital lobe<br>frontal lobe | 1.36 ± 0.05 (occipital)<br>0.93 ± 0.04 (frontal) | 9.4 T, indirect |
| This study | frontal lobe | 0.66 ± 0.07 | 7T, direct and indirect |
